# Access to routinely collected health data for clinical trials – review of successful data requests to UK registries

**DOI:** 10.1101/2020.04.08.20033373

**Authors:** Sarah Lensen, Archie Macnair, Sharon B Love, Victoria Yorke-Edwards, Nurulamin M Noor, Meredith Martyn, Alexandra Blenkinsop, Carlos Diaz-Montana, Graham Powell, Elizabeth Williamson, James Carpenter, Matthew R Sydes

**Affiliations:** MRC CTU at UCL WC1V 6LJ, Health Data Research UK; MRC CTU at UCL WC1V 6LJ; Department of Molecular and Clinical Pharmacology, University of Liverpool, L69 3BX; London School of Hygiene and Tropical Medicine WC1E 7HT; Medical Statistics, London School of Hygiene and Tropical Medicine WC1E 7HT

**Author notes:** ***Correspondence to:*** Sharon Love. These authors contributed equally (James Carpenter, Matthew R Sydes).

**Keywords:** Systematic review, Routinely-collected health data, registry, RCT

## Abstract

**Background:** Clinical trials generally each collect their own data despite routinely-collected health data (RCHD) increasing in quality and breadth. Our aim is to quantify UK-based randomised controlled trials (RCTs) accessing RCHD for participant data, characterise how these data are used and thereby recommend how more trials could use RCHD.

**Methods:** We conducted a systematic review of RCTs accessing RCHD from at least one registry in the UK between 2013-2018, for the purposes of informing or supplementing participant data. A list of all registries holding RCHD in the UK was compiled. In cases where registries published release registers, these were searched for RCTs accessing RCHD. Where no release register was available, registries were contacted to request a list of RCTs. For each identified RCT, information was collected from all publicly available sources (release registers, websites, protocol etc.). The search and data extraction was undertaken between Jan-2019 and May-2019.

**Results:** We identified 160 RCTs accessing RCHD between 2013 and 2018 from a total of 22 registries; this corresponds to only a very small proportion of all UK RCTs (approximately 3%). RCTs accessing RCHD were generally large (median sample size 1590), commonly evaluating treatments for cancer or cardiovascular disease. Most of the included RCTs accessed RCHD from NHS Digital (68%), and the most frequently accessed datasets were mortality (76%) and hospital visits (55%). RCHD was used to inform the primary trial (82%) and long-term follow-up (57%). There was substantial variation in how RCTs used RCHD to inform participant outcome measures. A limitation was the lack of information and transparency from registries and RCTs with respect to which datasets have been accessed and for what purposes.

**Conclusions:** In the last five years, only a small minority of UK-based RCTs have accessed RCHD to inform participant data. We ask for improved accessibility, confirmed data quality and joined up thinking between the registries and the regulatory authorities.

**Registration:** PROSPERO CRD42019123088

## BACKGROUND

Randomised controlled trials (RCTs) are the gold-standard method for evaluating healthcare interventions, and their results impact on policy, practice, and patient care. Substantial resources are dedicated to collection of trial data and participant follow-up. Consequently the costs of conducting large trials are substantial, maybe in the order of millions of pounds (1). However many national databases and registries collect data which map to common important healthcare events such as hospital admission, cancer registration and death. Use of this routinely-collected health data (RCHD) to replace or supplement traditional data capture should reduce trial costs, enabling a greater number of large, definitive trials and efficient long-term assessment of healthcare interventions.

This explains why the use of RCHD in RCTs has been labelled as a disruptive technology i.e. a technology which transforms current practice (2). A model exemplar is the TASTE trial, which randomised 7244 participants in two years within national Swedish registries, collected participant data from registries and yielded high impact results at a fraction of the cost of traditional RCTs (USD $300,000 or $50 per patient)(3, 4). The UK holds a large number of rich health datasets, linkable through a unique National Health Service (NHS) number. The availability of these datasets is growing, as are the technological capabilities of processing and storing this data. In response to this, Health Data Research (HDR) UK was established with the ambition to unleash the potential of RCHD to deliver “Better, Faster and More Efficient Trials” (5).

However, while RCHD is already being harnessed to enhance UK RCTs, anecdotal evidence suggests substantial barriers remain. Therefore, this systematic review set out to identify and characterise RCTs accessing RCHD in the UK to inform participant data, to describe how RCTs use these data, and to prioritise issues which need to be addressed

## METHODS

A systematic review of RCTs which have accessed RCHD to inform or supplement trial data.

### Eligibility

RCHD was defined as data which are collected for “administrative and clinical purposes without specific a priori research goals” (6). This included large, national, administrative resources (e.g. NHS Digital), national disease and healthcare audits and registries in each UK devolved nation (e.g. the National Emergency Laparotomy Audit). Hereafter we refer to these collectively as registries. Cohort studies, biobanks, NHS Safe Havens and electronic health records held only at the point of care, such as primary care records held within general practitioner (GP) practices, were excluded.

Eligible RCTs received RCHD from a registry between 2013 and 2018. This time-frame was selected to broadly align with the initiation of release registers in large national databases following the 2014 Partridge Review (7). For each included RCT, any additional access to RCHD from another registry and any previous access of RCHD prior to 2013 was also captured.

Eligible RCTs were those which accessed RCHD to inform either baseline or outcome measure data of trial participants. For at least one outcome measure, RCHD must have been used for any combination of: (i) replacing conventionally collected trial data; (ii) cross-checking against existing trial data (including participant-reported data); (iii) cross-checking RCHD from different sources; (iv) triggering the trial team to further investigate a possible outcome measure or event; (v) cost-effectiveness analysis and (vi) solely methodological purposes. This was captured separately for (a) the primary reporting period of the trial (i.e. baseline data or an outcome measure within the follow-up for capturing the primary trial outcome measure) and (b) for long-term follow-up.

We excluded RCTs if the RCHD was only accessed to plan or facilitate recruitment, e.g. to contact patients with an invitation of RCT enrolment, or to extrapolate results of RCTs to broader populations. The Protocol for this review was registered with PROSPERO at the stage of screening and data collection (CRD42019123088, registered 20 Feb 2019).

### RCT identification

First, we compiled a list of registries (healthcare databases, registries and audits) in the UK through internet searching, the Health Quality Improvement Program (HQIP) directory (8), contact with government and contracted organisations, and existing knowledge of UK registries (more information on registries approached in additional file [additional file 1]). Release registers were identified where possible; these are lists of all data released from a given registry, often including the purpose for which the data will be used and the specific datasets accessed. Where these were not available, registries were contacted to request a list of RCTs to which they had released RCHD.

Release registers from each source were de-duplicated prior to screening (to remove multiple instances of data releases for the same RCT from an individual registry). The resultant list was then searched for eligible RCTs by filtering for entries containing one or more of the following terms: rand*, trial, RCT, study, placebo, phase. The search results were then screened independently for potentially eligible RCTs by two authors. Disagreements were resolved by discussion and re-checking.

### Data collection and analysis

For each RCT identified, we sought information from within the release registers (e.g. ‘purpose statements’), RCT websites (including privacy statements, publications, protocols, statistical analysis plans, patient information sheets and consent forms), and other available sources including trial registration information. Publications for each RCT were searched for in major dissemination databases (e.g. MEDLINE, Google Scholar). More information about data collection is given in an additional file [additional file 4].

Data collection included information about the RCT (e.g. disease category, recruitment and publication status, primary outcome measure), the registry (e.g. NHS Digital), the RCHD accessed (e.g. Hospital Episode Statistics), and the way in which the data were used (e.g. linkage identifiers used, category of data use). Due to the large number of RCTs identified, we focussed more detailed data collection of information on the datasets accessed and the way in which the data were used to RCTs accessing RCHD between 2017 and 2018. Two authors independently extracted data onto a piloted data extraction form and any disagreements were resolved by discussion and re-checking. Data were subsequently entered into a clinical data management system (Elsevier’s MACRO (9)) and descriptive analyses were undertaken in Stata (version 15.1)(10). Trial teams were not contacted for information or clarification.

To enable a broad comparison of this cohort of RCTs with those conducted in the UK, we compared the descriptive characteristics of these RCTs with those reported in a recent cross-sectional analysis of UK Health Research Authority (HRA) approved RCTs (11).

### Patient and public involvement

No patients were involved in any component of the design, production, analysis, interpretation or writing up of the results of this review. We plan to disseminate the final results to the HDR UK Public Advisory Board, and request they disseminate the manuscript within their network as appropriate.

## RESULTS

### Results of the search

The search and extraction of data was undertaken between Jan-2019 and May-2019. 74 UK registries holding RCHD were identified, of which 13 maintained accessible release registers (Figure 1).

**Figure 1.**
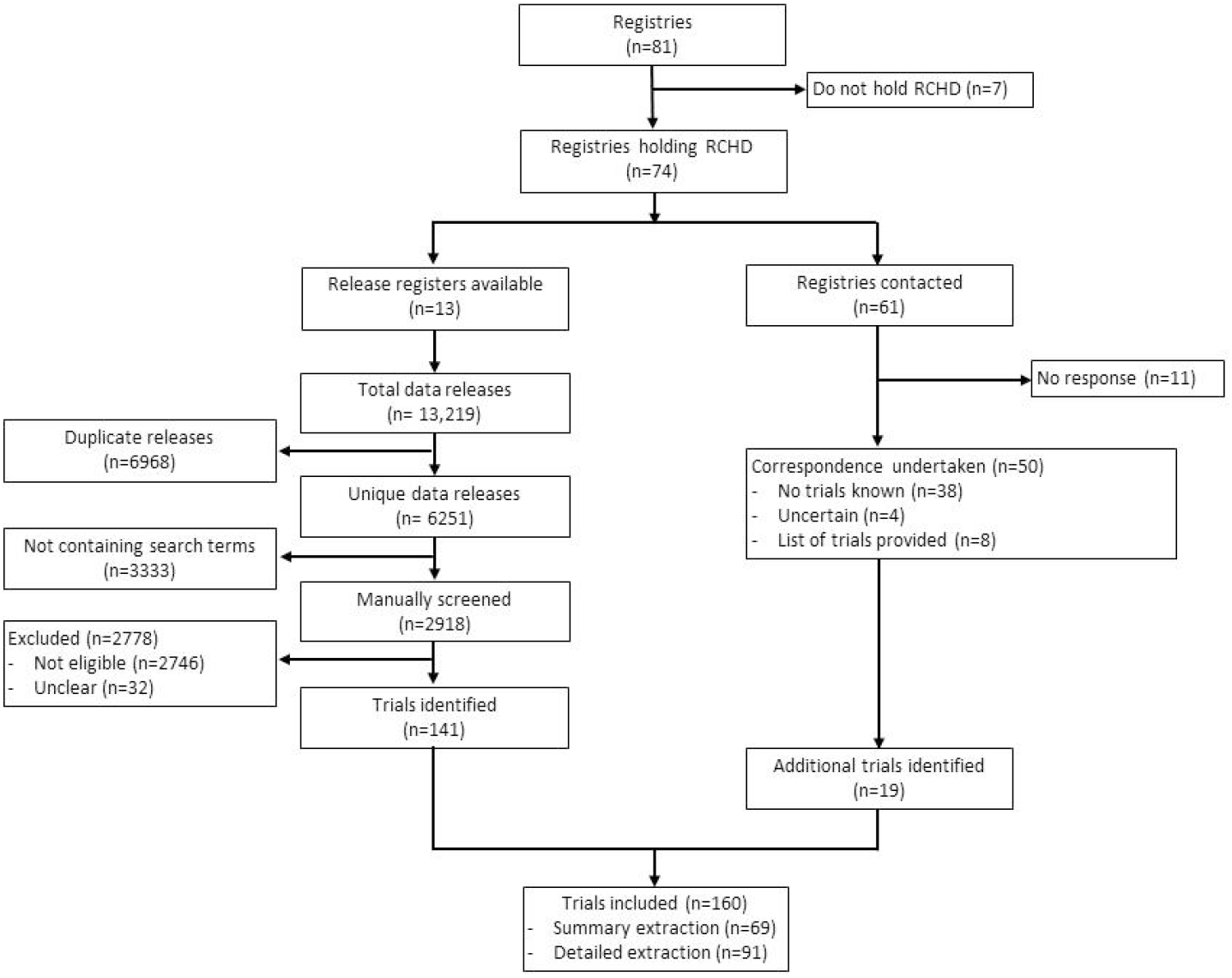
Identification of trials from registries. Each trial is only counted once. For instance, trials identified through both release register searches and notification by registries are only captured once. Of 13 registries with release registers available, 10 published comprehensive release registers and 3 provided a brief lists of projects receiving RCHD on the website.

These release registers listed more than 6,000 unique data releases. There were 2,918 releases identified in the search. These were manually screened and 141 RCTs were identified; corresponding to 2% (141/6251) of the total releases. The remaining 61 registries were contacted to request information about RCTs having accessed RCHD, resulting in a further 19 RCTs identified from eight registries. During the data extraction, we discovered one trial that had received data from one of the registries which had not otherwise provided a list of trials. This gave a total of 160 RCTs accessing RCHD from 22 registries between 2013 and 2018 (Figure 1). Although all RCTs had accessed RCHD between 2013 and 2018, they were conducted in varying time periods, with recruitment start dates ranging from 1979 to 2018. Detailed data collection, for trials accessing data in 2017-18, involved 91/160 trials.

### RCT characteristics

The 160 included RCTs were generally large (median sample size 1590, range 41 – 6,000,000), although 11% (17/160) described themselves as pilot or feasibility trials (Table 1). The majority (85%, 136/160) were individually randomised trials and 15% (24/160) were cluster randomised. The most common disease categories were cancer (29%, 47/160) and cardiovascular disease (29%, 46/160), and the most common primary outcome measure was survival/death (45%, 72/160). Only 20% (32/160) of the RCTs were international, recruiting at additional sites outside of the UK. A small number of RCTs had publications available which included outcome measures informed by access to RCHD. Of these, 83% (29/35) had one or more results publication in a high profile medical journal.

**Table 1.**
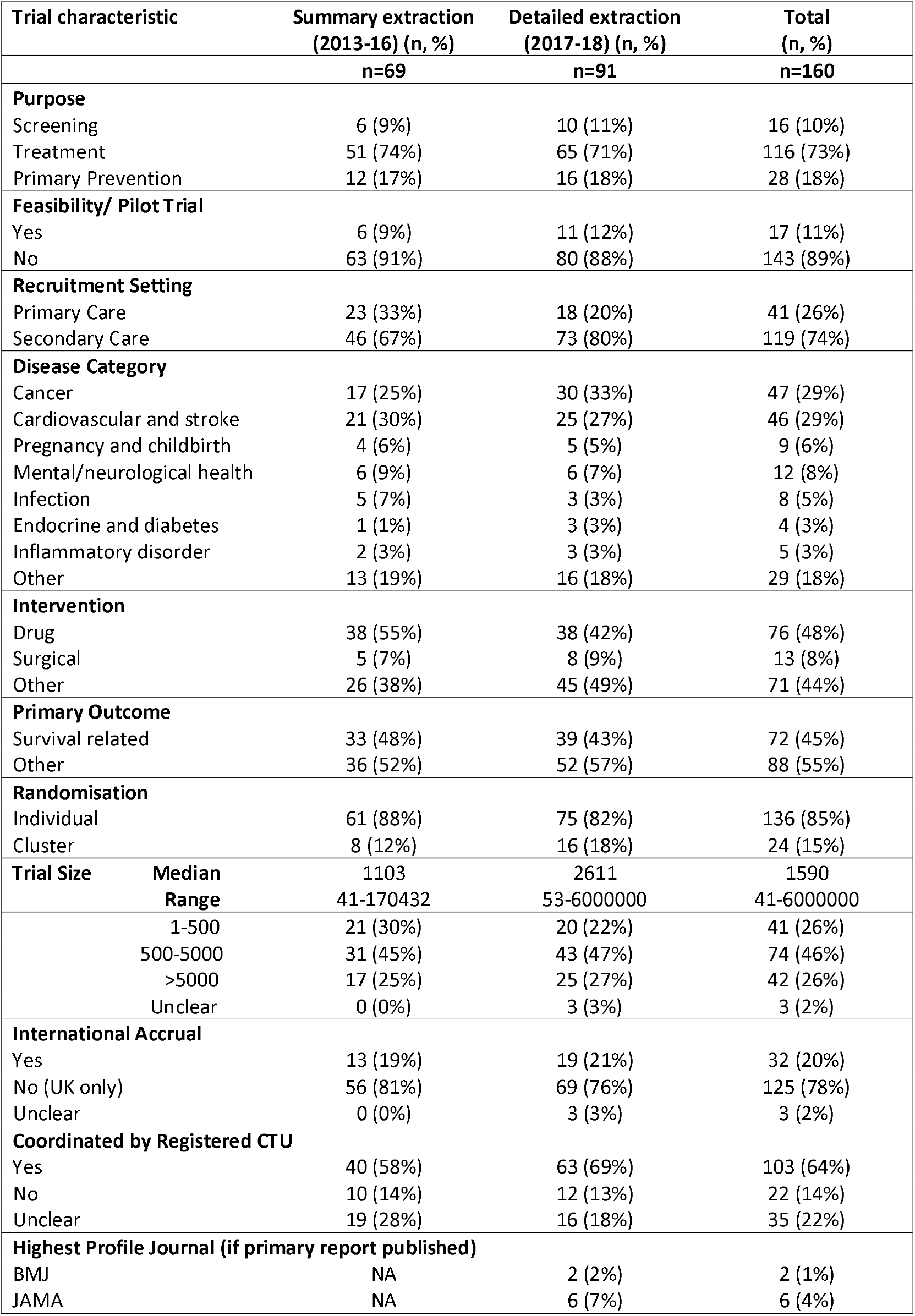
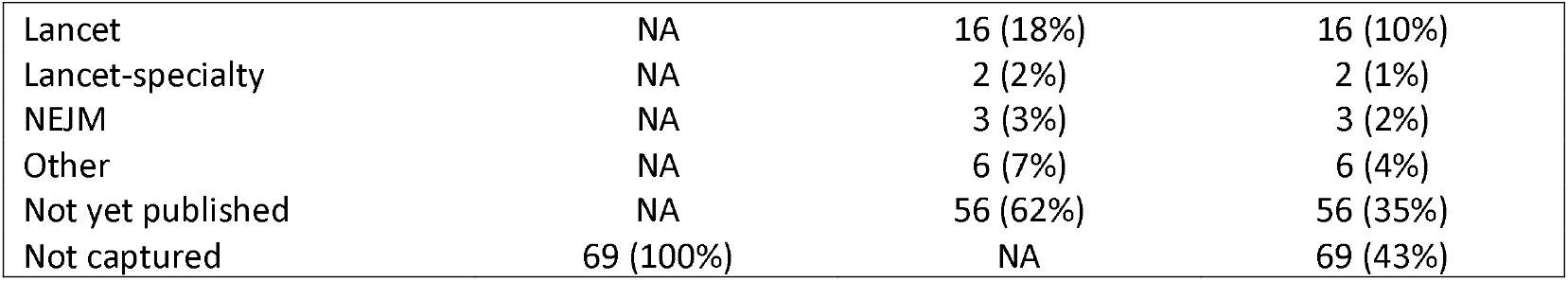
Trial characteristics.

| Trial characteristic |  | Summary extraction<br>(2013-16) (n, %) | Detailed extraction<br>(2017-18) (n, %) | Total<br>(n, %) |
| --- | --- | --- | --- | --- |
|  |  | n=69 | n=91 | n=160 |
| **Purpose** |  |  |  |  |
| Screening |  | 6 (9%) | 10 (11%) | 16 (10%) |
| Treatment |  | 51 (74%) | 65 (71%) | 116 (73%) |
| Primary Prevention |  | 12 (17%) | 16 (18%) | 28 (18%) |
| **Feasibility/ Pilot Trial** |  |  |  |  |
| Yes |  | 6 (9%) | 11 (12%) | 17 (11%) |
| No |  | 63 (91%) | 80 (88%) | 143 (89%) |
| **Recruitment Setting** |  |  |  |  |
| Primary Care |  | 23 (33%) | 18 (20%) | 41 (26%) |
| Secondary Care |  | 46 (67%) | 73 (80%) | 119 (74%) |
| **Disease Category** |  |  |  |  |
| Cancer |  | 17 (25%) | 30 (33%) | 47 (29%) |
| Cardiovascular and stroke |  | 21 (30%) | 25 (27%) | 46 (29%) |
| Pregnancy and childbirth |  | 4 (6%) | 5 (5%) | 9 (6%) |
| Mental/neurological health |  | 6 (9%) | 6 (7%) | 12 (8%) |
| Infection |  | 5 (7%) | 3 (3%) | 8 (5%) |
| Endocrine and diabetes |  | 1 (1%) | 3 (3%) | 4 (3%) |
| Inflammatory disorder |  | 2 (3%) | 3 (3%) | 5 (3%) |
| Other |  | 13 (19%) | 16 (18%) | 29 (18%) |
| **Intervention** |  |  |  |  |
| Drug |  | 38 (55%) | 38 (42%) | 76 (48%) |
| Surgical |  | 5 (7%) | 8 (9%) | 13 (8%) |
| Other |  | 26 (38%) | 45 (49%) | 71 (44%) |
| **Primary Outcome** |  |  |  |  |
| Survival related |  | 33 (48%) | 39 (43%) | 72 (45%) |
| Other |  | 36 (52%) | 52 (57%) | 88 (55%) |
| **Randomisation** |  |  |  |  |
| Individual |  | 61 (88%) | 75 (82%) | 136 (85%) |
| Cluster |  | 8 (12%) | 16 (18%) | 24 (15%) |
| **Trial Size** | **Median** | 1103 | 2611 | 1590 |
|  | **Range** | 41-170432 | 53-6000000 | 41-6000000 |
|  | 1-500 | 21 (30%) | 20 (22%) | 41 (26%) |
|  | 500-5000 | 31 (45%) | 43 (47%) | 74 (46%) |
|  | >5000 | 17 (25%) | 25 (27%) | 42 (26%) |
|  | Unclear | 0 (0%) | 3 (3%) | 3 (2%) |
| **International Accrual** |  |  |  |  |
| Yes |  | 13 (19%) | 19 (21%) | 32 (20%) |
| No (UK only) |  | 56 (81%) | 69 (76%) | 125 (78%) |
| Unclear |  | 0 (0%) | 3 (3%) | 3 (2%) |
| **Coordinated by Registered CTU** |  |  |  |  |
| Yes |  | 40 (58%) | 63 (69%) | 103 (64%) |
| No |  | 10 (14%) | 12 (13%) | 22 (14%) |
| Unclear |  | 19 (28%) | 16 (18%) | 35 (22%) |
| **Highest Profile Journal (if primary report published)** |  |  |  |  |
| BMJ |  | NA | 2 (2%) | 2 (1%) |
| JAMA |  | NA | 6 (7%) | 6 (4%) |

|  |  |  |  |
| --- | --- | --- | --- |
| Lancet | NA | 16 (18%) | 16 (10%) |
| Lancet-specialty | NA | 2 (2%) | 2 (1%) |
| NEJM | NA | 3 (3%) | 3 (2%) |
| Other | NA | 6 (7%) | 6 (4%) |
| Not yet published | NA | 56 (62%) | 56 (35%) |
| Not captured | 69 (100%) | NA | 69 (43%) |

The majority of RCTs were clearly coordinated through a UK Clinical Research Collaboration (UKCRC) Registered Clinical Trials Unit (64%, 103/160 coordinated by a registered CTU, 14%, 22/160 were not, 22%, 35/160 were unclear) (Table 1). Of all 51 currently registered CTUs, 63% (32/51) had accessed RCHD for at least one RCT in this cohort. Of these CTUs, the median number of RCTs from this cohort was 2 (range 1-11).

RCTs accessing RCHD were more often conducted in cancer and cardiovascular populations compared to RCTs submitted for an ethical opinion via the HRA in 2015 (29% vs 10%, and 29% vs 17%, respectively), were more likely to recruit from primary care settings (26% vs 5%), to be based only in the UK (78% vs 50%) and to be cluster-randomised (15% vs 3%). RCHD RCTs had larger sample sizes on average (median 1590 vs 275) than those submitted to the HRA (Table 2). RCTs accessing RCHD were less likely to be feasibility/pilot studies (11% vs 18%). We identified 160 trials accessing RCHD over a five-year period (32 trials per year), which is equivalent to approximately 3% (32/963) of all RCTs approved by the HRA in 2015.

**Table 2.**
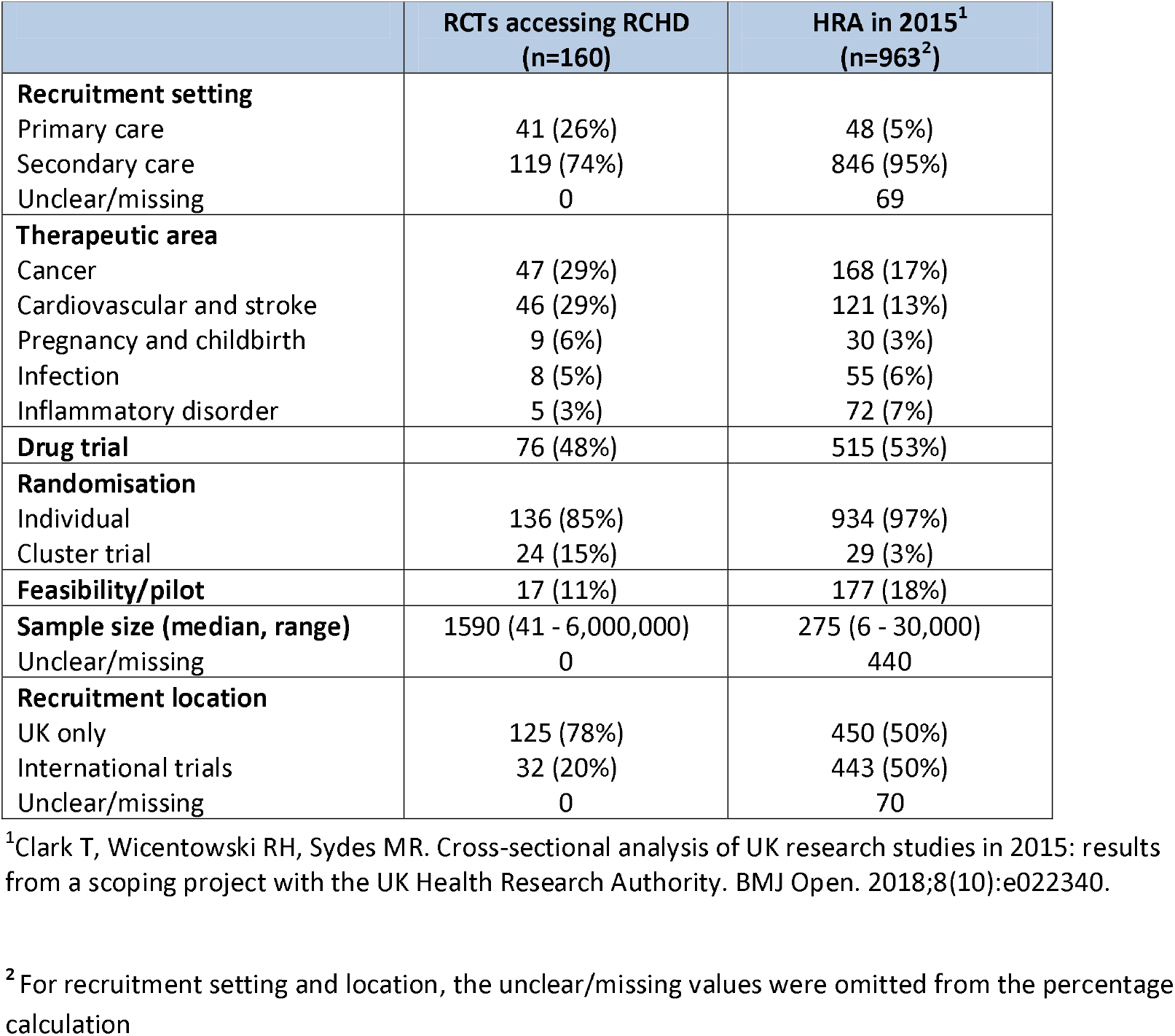
Comparison of RCTs accessing RCHD with trials evaluated by the HRA in 2015. This table only includes data fields which were comparable between the two sources. Sample size targets in the HRA cohort are limited to those not described as phase I/II trials. Data obtained from Clark et al 2018, including unpublished supplementary appendices (1).

|  | **RCTs accessing RCHD**<br>**(n=160)** | **HRA in 2015<sup>1</sup>**<br>**(n=963<sup>2</sup>)** |
| --- | --- | --- |
| **Recruitment setting** |  |  |
| Primary care | 41 (26%) | 48 (5%) |
| Secondary care | 119 (74%) | 846 (95%) |
| Unclear/missing | 0 | 69 |
| **Therapeutic area** |  |  |
| Cancer | 47 (29%) | 168 (17%) |
| Cardiovascular and stroke | 46 (29%) | 121 (13%) |
| Pregnancy and childbirth | 9 (6%) | 30 (3%) |
| Infection | 8 (5%) | 55 (6%) |
| Inflammatory disorder | 5 (3%) | 72 (7%) |
| **Drug trial** | 76 (48%) | 515 (53%) |
| **Randomisation** |  |  |
| Individual | 136 (85%) | 934 (97%) |
| Cluster trial | 24 (15%) | 29 (3%) |
| **Feasibility/pilot** | 17 (11%) | 177 (18%) |
| **Sample size (median, range)** | 1590 (41 - 6,000,000) | 275 (6 - 30,000) |
| Unclear/missing | 0 | 440 |
| **Recruitment location** |  |  |
| UK only | 125 (78%) | 450 (50%) |
| International trials | 32 (20%) | 443 (50%) |
| Unclear/missing | 0 | 70 |

<sup>1</sup>Clark T, Wicentowski RH, Sydes MR. Cross-sectional analysis of UK research studies in 2015: results from a scoping project with the UK Health Research Authority. BMJ Open. 2018;8(10):e022340.
<sup>2</sup>For recruitment setting and location, the unclear/missing values were omitted from the percentage calculation

### RCHD access and use

NHS Digital was by far the most commonly accessed registry: 68% (108/160) trials accessed RCHD from NHS Digital (Table 3).

**Table 3.** Registries and datasets accessed. Registries accessed was captured for all 160 trials. Information about datasets accessed from these registries was captured only for those 91 accessing RCHD between 2017 and 2018. The fields are not mutually exclusive as one trial may have accessed data from multiple registries; and multiple datasets can be accessed via a single registry. Percentages are calculated using the column header denominators. Hospital visits includes all Hospital Episode Statistics (Outpatient, Inpatient, Accident and Emergency, and Critical Care), Patient Episode Database for Wales (PEDW), and Scottish Morbidity Records (SMR). Acronyms: Information Services Division (ISD), Public Health England (PHE), Secure Anonymised Information Linkage (SAIL), Intensive Care National Audit & Research Centre (ICNARC), NHS Wales Informatics Service (NWIS), Paediatric Intensive Care Audit Network (PICANet), Clinical Practice Research Datalink (CPRD), NHS Blood and Transplant (NHSBT), Trauma Audit and Research Network - Major Trauma Audit (TARN), National Emergency Laparotomy Audit (NELA), Neonatal Research Database (NNRD), Public Health Wales (PHW), UK Renal Registry (UKRR), Department of Health (DOH), Falls and Fragility Fractures Audit programme (FFFAP), Honest Broker Service, Northern Ireland Statistics and Research Agency (HBS), National Institute for Cardiovascular Outcomes Research (NICOR), Northern Ireland Cancer Registry (NICR), Out-of-Hospital Cardiac Arrest Outcomes (OHCAO) Registry, UK Cystic Fibrosis Registry (UKCFR)

| Registry | Total Trials<br>n=160 | Total Trials<br>(2017-2018)<br>n=91 | Datasets accessed<br>Total Trials (2017-2018) n=91 |  |  |  |
| --- | --- | --- | --- | --- | --- | --- |
|  |  |  | Death<br>n=69 | Hospital<br>visits n=50 | Cancer<br>registration<br>n=29 | Other n=26 |
| **NHS-Digital** | 108 (68%) | 59 (65%) | 58 (84%) | 34 (68%) | 22 (76%) | 4 (15%) |
| **ISD-Scotland** | 35 (22%) | 25 (27%) | 16 (23%) | 13 (26%) | 7 (24%) | 2 (8%) |
| **PHE** | 15 (9%) | 11 (12%) | 3 (4%) | 6 (12%) | 10 (34%) | 1 (4%) |
| **SAIL** | 9 (6%) | 6 (7%) | 2 (3%) | 5 (10%) | 2 (7%) | 1 (4%) |
| **ICNARC** | 7 (4%) | 4 (4%) | 1 (1%) | 0 (0%) | 0 (0%) | 4 (15%) |
| **NWIS** | 7 (4%) | 6 (7%) | 1 (1%) | 6 (12%) | 1 (3%) | 0 (0%) |
| **PICANet** | 6 (4%) | 2 (2%) | 0 (0%) | 0 (0%) | 0 (0%) | 2 (8%) |
| **CPRD** | 4 (3%) | 1 (1%) | 0 (0%) | 1 (2%) | 0 (0%) | 1 (4%) |
| **NHSBT** | 3 (2%) | 3 (3%) | 0 (0%) | 0 (0%) | 0 (0%) | 2 (8%) |
| **TARN** | 3 (2%) | 2 (2%) | 1 (1%) | 1 (2%) | 0 (0%) | 2 (8%) |
| **NELA** | 2 (1%) | 2 (2%) | 0 (0%) | 0 (0%) | 0 (0%) | 2 (8%) |
| **NNRD** | 2 (1%) | 2 (2%) | 1 (1%) | 1 (2%) | 0 (0%) | 2 (8%) |
| **PHW** | 2 (1%) | 1 (1%) | 0 (0%) | 0 (0%) | 0 (0%) | 1 (4%) |
| **UKRR** | 2 (1%) | 2 (2%) | 0 (0%) | 1 (2%) | 1 (3%) | 0 (0%) |
| **ResearchOne** | 2 (1%) | 2 (2%) | 0 (0%) | 0 (0%) | 0 (0%) | 2 (8%) |
| **DOH** | 1 (1%) | 0 (0%) | 0 (0%) | 0 (0%) | 0 (0%) | 0 (0%) |
| **FFFAP** | 1 (1%) | 1 (1%) | 0 (0%) | 0 (0%) | 0 (0%) | 1 (4%) |
| **HBS** | 1 (1%) | 1 (1%) | 0 (0%) | 0 (0%) | 0 (0%) | 1 (4%) |
| **NICOR** | 1 (1%) | 1 (1%) | 1 (1%) | 0 (0%) | 0 (0%) | 0 (0%) |
| **NICR** | 1 (1%) | 1 (1%) | 0 (0%) | 0 (0%) | 1 (3%) | 0 (0%) |
| **OHCAO** | 1 (1%) | 1 (1%) | 0 (0%) | 0 (0%) | 0 (0%) | 1 (4%) |
| **UKCFR** | 1 (1%) | 1 (1%) | 0 (0%) | 0 (0%) | 0 (0%) | 1 (4%) |

The second most common was Information Services Division in Scotland 22%, 35/160). Most of the RCTs accessed RCHD from one registry (79%, 126/160); 14% (22/160) accessed data from two registries, 5% (8/160) from 3 registries, and 3% (4/160) from 4 or more. A small number of RCTs were completely embedded (i.e. participants were recruited from and followed-up) in the registry (12%, 11/91).

Of the 160 RCTs, 91 had received a total of 134 data releases in the years 2017-2018 and were selected for detailed data extraction. Identifiers used for linkage were often unclear (46%, 62/134), however when assessable, the most common fields were NHS Number (94%, 68/72), date of birth (85%, 61/72) and participant name (56%, 40/72) (an additional file shows Table S2 [additional file 2]). The most common datasets accessed were mortality (76%, 69/91), hospital visits (55%, 50/91) and cancer registration (32%, 29/91) (Table 3). Almost half of the included RCTs (47%, 43/91) accessed RCHD to inform the primary trial outcome measure. Of RCTs using RCHD only for at least one outcome measure, 38% (20/52) were drug trials.

36 out of 91 RCTs (40%, 36/91) accessed RCHD for both the primary and long-term follow-up (Table 4); 21% (19/91) of RCTs accessed one or more RCHD only for long-term follow-up and 45% (41/91) accessed one or more RCHD exclusively for the primary with no obvious planned long-term follow-up.

**Table 4.** Categories describing how RCHD was used to inform or supplement participant data. These categories were developed for the purpose of this review, and are not mutually exclusive. For example, RCTs may use RCHD for both cross-checking against existing trial data and against a second source of RCHD. Additionally, RCTs may use RCHD from multiple sources in different ways. Percentages are calculated using the column header denominators.

| **Data use category** | **Description** | **Example** | **All trials**<br>**N=91** | **Primary**<br>**N=74** | **Long-term**<br>**follow-up**<br>**N=52** | **Both**<br>**N=36** |
| --- | --- | --- | --- | --- | --- | --- |
| **1** | The RCHD alone is used as trial data, with no cross-checking or comparison against any other data | One of the included RCTs was a cluster-randomised trial of GP practices. Outcome data specific to the trial were extracted at the level of the cluster (GP practice) from CPRD. | 52 (57%) | 33 (45%) | 34 (65%) | 15 (42%) |
| **2** | RCHD is used to cross-check against or verify known trial data i.e. data already being collected on CRFs as part of the trial (arising from clinical observations and measurements) | In one case, death and cause of death of participants are captured on CRFs at each site. NHS-Digital also send the trial team quarterly reports of all new deaths. The trial team then compare the events and cause of death from both sources. | 27 (30%) | 24 (32%) | 6 (12%) | 3 (8%) |
| **3** | RCHD is used to cross-check against or verify self-reported trial data i.e. data already being collected from participants, for example by questionnaire | In one trial, participants are asked to complete a questionnaire every 3 months which asks if they had any unexpected stays in hospital. This information is cross-checked against Hospital Episode Statistics data obtained from NHS-Digital. | 28 (31%) | 22 (30%) | 11 (21%) | 5 (14%) |
| **4** | RCHD is used to alert or flag trial teams to a potential outcome/event, prompting medical note review to confirm the outcome/event. The specific outcome/events being flagged are not otherwise being captured as trial data. This may be accompanied by clinical end-point review or adjudication of events and outcomes. | In one RCT, participants were flagged in the UK Transplant Registry for notification of transplant rejection and failure. When participants were identified as having a transplant rejection or failure, study staff sought extra information from hospital records. The collated information was redacted and used for central adjudication by trained clinicians. | 22 (24%) | 19 (26%) | 9 (17%) | 6 (17%) |
| **5** | RCHD from one source is used to cross-check against or compared with RCHD from another source | One of the included RCTs accessed RCHD from NHS-Digital, ICNARC and OHCAO, and all had provided the same fields - such as length of stay in intensive care. | 9 (10%) | 7 (9%) | 6 (12%) | 4 (11%) |
| **6** | RCHD is being used for health economic analysis or cost-effectiveness purposes, rather than a clinical outcome | In one study, Hospital Episode Statistics data was used to calculate the cost of secondary resource use within 90 days of randomisation | 25 (27%) | 21 (28%) | 12 (23%) | 8 (22%) |
| **7** | RCHD is not used directly for trial purposes, but to evaluate the quality of these data compared to trial data or other RCHD, or RCHD is used to generate an algorithm or equation that hopes to predict or replicate the frequency of events/outcomes. | In a breast cancer trial, cancer data from NCRAS were accessed and compared against the trial data to assess the completeness, validity, and consistency of the two data sources. | 11 (12%) | 11 (15%) | 1 (2%) | 1 (3%) |
| **Unclear** | - | - | 13 (14%) | 9 (12%) | 9 (17%) | 5 (14%) |

Most commonly, RCHD alone was used for at least one trial outcome measure (57%, 52/91) (Table 4). One third of RCTs used RCHD for cross-checking, either of trial data (30%, 27/91) or participant-reported data (31%, 28/91). Use of RCHD to trigger case review was also common (24%, 22/91), as was use of the data to conduct cost-effectiveness analysis (27%, 25/91). Use of RCHD for methodological reasons was uncommon (12%, 11/91), as was release for comparison of two or more RCHD sources (10%, 9/91). RCTs using RCHD for long-term follow-up were more likely to use RCHD alone to inform outcome measures, and less likely to conduct cross-checking against trial or participant-reported data, or to use the data for methodological purposes. Overall, there was substantial variation in how trials used RCHD to inform participant outcome measures. For example, among the 74 trials using RCHD within the primary reporting period, 37 different combinations of data use were captured (an additional file shows Table S3 [additional file 3]). Among the 36 RCTs using RCHD for the primary report and long-term follow-up, 56% (20/36) used the data differently for these two stages of the study for at least one outcome measure, for example shifting from cross-checking of trial data for the primary reporting to RCHD only during the long-term follow-up.

## DISCUSSION

The increase in the scope, accessibility, and richness of RCHD presents an unprecedented opportunity for better health research(12). Use of RCHD for trial outcome measures may be a cost-effective means of obtaining data, limiting the burden on trial staff and participants in attending for trial visits or replying to questionnaires, especially for longer-term data collection. Use of RCHD may also minimise attrition in RCTs where datasets have national coverage, reduce issues of self-reported outcome measures which are prone to recall bias (e.g. recalling diagnoses or operations from hospital visits), and could limit ascertainment bias where the clinicians and coders are not aware of trial participation. However, are RCHD replacing case report forms in clinical trials and, if not, why not?

To the best of our knowledge, this is the first review to summarise the accessing of RCHD by randomised trials in the UK by reviewing the sources of data, and the first to assess the use of these data specifically for trial outcome measure assessment. We identified 160 trials accessing RCHD to inform participant data from 22 registries in the UK between 2013 and 2018, with many (47%, 43/91) using it for the primary outcome measure. This corresponds to approximately 32 trials a year, which is about 3% of the trials seeking HRA approval annually (11). Alongside this, RCTs accessing RCHD accounted for only 2% of the data releases from included registries. Since most trial patient data are captured in the hospital records, this suggests the potential of RCHD in trials is largely untapped.

We observed considerable variation in the use of RCHD, most commonly to inform or supplement outcome measures in primary trial report and long-term follow-up. Only 52/91 (57%) used RCHD alone for the collection of at least one trial outcome measure i.e. even when used, the data are duplicated from trial-specific sources – further evidence that the potential of RCHD is largely unrealised.

Only a very small proportion of UK trials appear to be successfully accessing RCHD. Our findings are consistent with anecdotal evidence that one barrier to greater access and use may be lack of awareness among trialists regarding the availability and potential utility of this information for trial follow-up. There is no national directory of registries which lists sources of RCHD available to researchers. The National Institute for Health Research (NIHR) Health Data Finder for Research contains only 18 datasets (18). Half of the registries identified for this review confirmed they had not provided data to RCTs and may represent an underutilised resource.

Both for us (as reviewers) and for trialists, the lack of a comprehensive list of RCHD registries and the data they hold, is a challenge. Further, the majority of registries we identified did not maintain a register of approved data releases. A number of release registers had brief information (e.g. only application titles), and some registries were unable to advise whether their RCHD had been released for this purpose. Therefore, our search may have missed eligible trials. For trialists, this makes it more difficult to keep abreast of how these data may be used, hindering the uptake of RCHD by the community. A further barrier is that many publications about the included trials which were expected to include RCHD, made no mention of it. So it was often not clear from publically-available sources exactly how RCHD would be used with a trial (note we deliberately did not contact trial teams for information or clarification, as our aim was to assess information which was publically available). The forthcoming CONSORT extension for RCTs using cohorts and accessing electronic health records should help to improve transparency in reporting (13) and enable the community to keep abreast of developments.

Other recent reviews in this area have summarised characteristics of trials in other settings, including those utilising these data for at least one trial outcome measure (14, 15), and for the long-term extension of completed trials (16,17). These reviews identified similar types of trials accessing RCHD, in terms of trial characteristics. However, due to the traditional literature searches employed by these reviews, they identified only a handful of the UK trials identified here; by reviewing release registers rather than publications we found more trials are receiving data than are mentioning it in their publications.

Reliance on data provision from registries raises unpredictable, and potentially extremely time consuming, challenges relating to data access and retention. For example, changes to registry names can render participant consent invalid if it no longer references the correct provider name. Individuals at registry organisations are also known to have provided contrary information on specific consent form wording (19). Many researchers report long delays in the application process, impacting on timely data collection and trial completion, with reports of RCTs being unable to publish trial results due to issues with data access (20), and in one RCT failure to gain access to mortality data has necessitated a change to the primary trial outcome measure (21). Cancer registration data, collected by Public Health England, have previously been available through NHS Digital, however provision of this data stopped for a period of more than two years. Such unscheduled lapses in data availability introduce substantial risk for RCTs relying on these cancer registration notifications through this route. One RCT reported failure by the registry to update flagging of new patients as recruitment continued: the trial team received death information only for the initial half of their cohort (22). Additionally, many registries do not permit ongoing retention or onward sharing of the datasets, creating conflict with key trial processes such as data archiving, data sharing and individual participant data meta-analysis (19, 23).

The administrative nature of some RCHD sources, the external coding and validation processes employed, and lack of oversight and visibility of data collection, processing, and audit trails raise concerning implications for Good Clinical Practice (GCP) adherence (24). The data used in clinical trials have to be the same as the source data to be GCP compliant. There are accounts of data quality issues from RCHD, even for clearly-objective outcome measures such as death (25-27) though cardiovascular outcomes seem more promising (28). A standardised, systematic approach to data quality assessment, ideally as a coordinated series of multi-RCT study-within-a-trial (SWAT), would provide empirical evidence of the quality of RCHD and traditional trial data.. Registry processes for data collection and editing would also need to be assessed.

The timeliness of RCHD is key. While primary care data, for example held by Clinical Practice Research Datalink (CPRD), can be extracted easily from multiple GP practices across software systems (Vision or EMIS), provision of secondary care data such as Hospital Episode Statistics generally has delayed capture, and is received in batch files every month or quarter. Certainly these data cannot be relied upon for the timely reporting of serious adverse events (e.g. requiring hospital admission).

## Conclusion

Only a tiny percentage of UK-based RCTs have accessed RCHD in the last five years to inform participant data, and few of these are exclusively relying on RCHD, despite the fact that most patient data are captured by hospital systems. Further, while most RCTs appear to be utilising similar datasets from a small number of registries, the way in which the RCHD is used to inform or supplement trial data appeared to vary substantially. Barriers to lack of utilisation include access to data and fitness of RCHD for research purposes.

Our review supports concerns that exploiting the potential of RCHD in trials is hindered (Table 5). Targeting resource to developing robust solutions to overcome these hurdles and enable a step change for clinical trials is urgently needed such that UK trials can fully harness the power of RCHD to conduct more efficient RCTs.

**Table 5:**
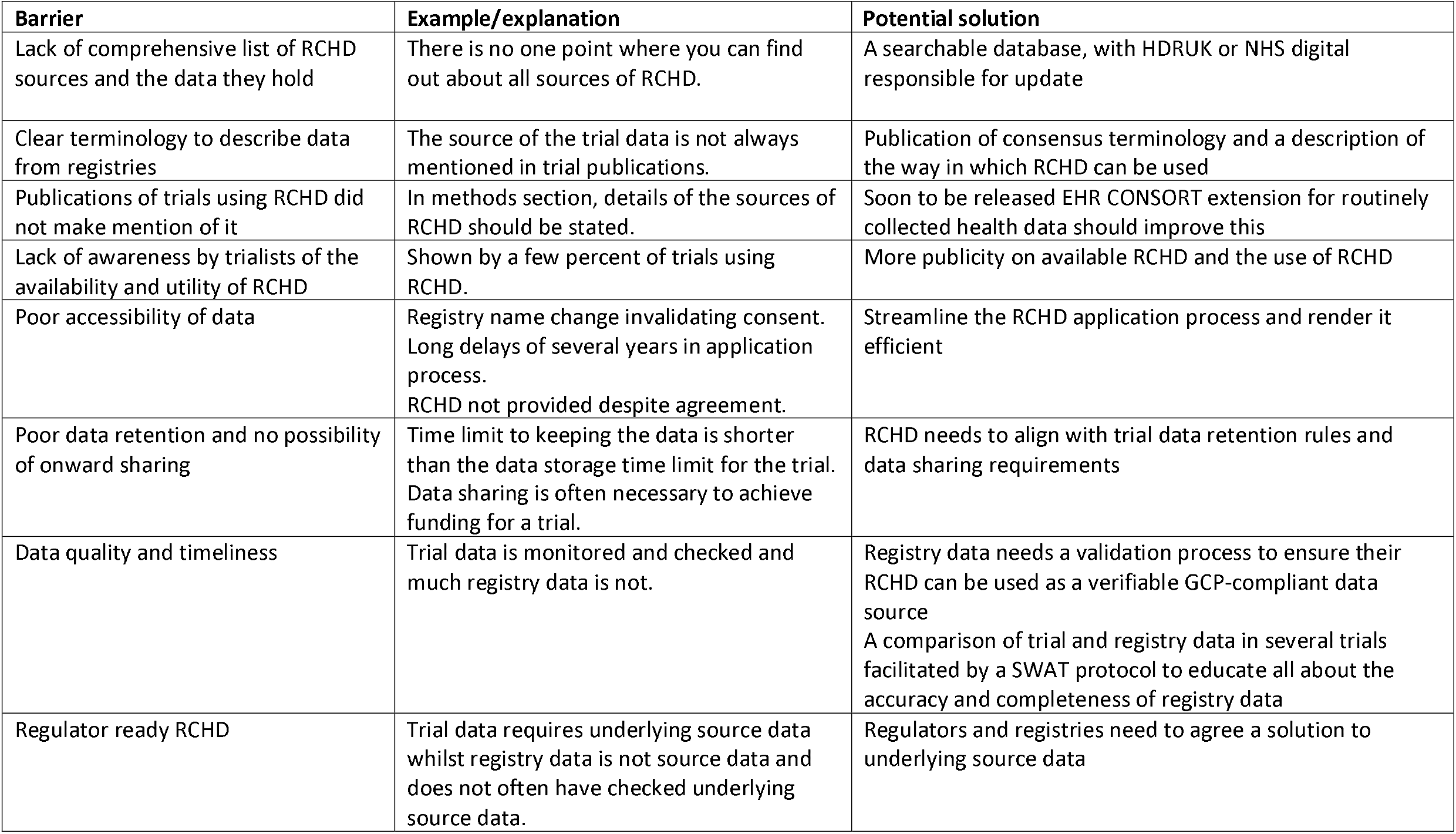
Barriers to use of routinely collected health data and potential solutions.

## Data Availability

All of the information is publicly available. The dataset and technical appendices are available upon request as per the controlled access approach of the MRC CTU at UCL. Please contact the corresponding author for more information

## List of abbreviations

HDR UK: Health Data Research UK
NIHR: National Institute for Health Research
RCT: randomised controlled trial
RCHD: routinely-collected health data
UKCRC: UK Clinical Research Collaboration

## Declarations

### Ethics approval and consent to participate

Not required as this was a review of RCTs accessing RCHD and no patient-level data were used

### Consent for publication

Not applicable

### Authors’ contributions

SLe conceived the idea for the review, drafted the protocol, and drafted the manuscript. SLe, VYE, SLo, MS, JC developed the protocol and planned the project. SLe, AM, VYE, SLo, EW, GP, JC and MS performed screening and RCT identification. SLe, AM, VYE, SLo, EW, GP, MS, NN, AB and MM performed data extraction. CDM lead the development of the computer application used for data collection. SLe, SLo and VYE performed the data analysis. All authors contributed to the paper writing and reviewed the last version of the manuscript.

#### Acknowledgements

Thank you to the UKCRC Registered CTU Network and to all those who completed the questionnaire.

## Authors’ information

SBL, VYE, SL and MS are part of the MRC CTU at UCL Trial Conduct Team. RCHD is one of the three streams of research from this team.

## Additional files

| Filename | File<br>format | title | description |
| --- | --- | --- | --- |
| Additional file_1 | docx | Table S1 | Registries searched<br>and approached |
| Additional file_2 | docx | Table S2 | Linkage identifier<br>combinations by<br>frequency of use<br>(detailed extraction<br>2017-2018) |
| Additional File_3 | docx | Table S3 | Data use<br>combinations by<br>frequency of use,<br>among trials using<br>data for the primary<br>report (n=74) |
| Additional file_4 | docx | Appendix 1 | Additional<br>information about<br>data collection |

